# Patient-reported benefit and speech perception in 24,570 hearing aid users: a nationwide study of the French Client-Oriented Scale of Improvement (COSI)

**DOI:** 10.64898/2026.11.12.26367934

**Authors:** Perrine Morvan, Marta Campi, Gareth W. Peters, Hung Thai-Van

## Abstract

**Objective:** To examine whether patient-reported benefit measured with the French Client-Oriented Scale of Improvement (COSI) is associated with speech perception, to describe its measurement properties, and to assess how well its predefined categories capture patients’ listening needs.

**Design:** Nationwide, retrospective, cross-sectional study of routinely collected clinical data (2024), with one questionnaire per patient.

**Study sample:** 24,570 adults fitted with hearing aids in 780 French hearing-care centres.

**Results:** Perceived benefit was only weakly correlated with post-fitting speech perception in quiet and in noise (*r* = *−*0.11 to *−*0.15). Perceived change was not, or only negligibly, related to measured change (*r* = 0.06 in quiet and 0.08 in noise). Age was moderately correlated with speech perception (*r* = 0.34) but not with perceived benefit (*r* = *−*0.05). Both COSI subscores showed a ceiling effect (35–40% of patients at the highest score) and agreed moderately (ICC = 0.72). One patient in four (24.5%) reported a first-ranked need outside the predefined categories, most often device discretion, tinnitus or cost.

**Conclusions:** In routine practice, COSI benefit was largely dissociated from speech perception. The COSI and speech tests should be used together, and the COSI categories should be updated.

## Introduction

Hearing loss (HL) affects communication, social participation and quality of life [1–3]. Despite these broad consequences, the clinical assessment and management of HL still rely mainly on behavioural tests: pure-tone audiometry to determine the severity of HL [4], and speech perception tests in quiet (SIQ) [5] and in noise (SIN) [6] to describe functional auditory capacity. These measures are essential for diagnosis, device selection and fitting, but they only partly reflect the listening difficulties that patients experience in daily life [7]. Patients with similar audiometric profiles often report very different levels of communication difficulty, participation restriction and hearing aid (HA) benefit [8–10], and speech perception scores do not consistently predict satisfaction or perceived improvement after rehabilitation [11]. Conventional audiological tests alone may therefore be insufficient to describe rehabilitation outcomes.

To address this gap, international organisations, including the World Health Organization (WHO) and American Speech-Language-Hearing Association (ASHA), recommend the use of patient-reported outcome measures (PROMs) in routine audiological care [12, 13]. PROMs give direct insight into the patient’s perspective and capture the functional, social and emotional consequences of HL that behavioural tests may not reflect [14, 15]. Open-ended questions can also improve the assessment of chronic conditions when they are used alongside PROMs [16]. However, as Saunders et al. [17] highlighted, choosing and administering appropriate outcome measures is complex: clinicians must use several tools with the same competence as diagnostic tests, and researchers must show their added value.

Among available PROMs, the Client Oriented Scale of Improvement (COSI) [18, 19] is commonly used in audiology. Unlike fixed-item questionnaires, the COSI asks patients to name, in their own words, the listening needs in which they most want to improve, and then to rate the benefit obtained in these needs. To allow comparisons between patients, these open-ended needs are usually classified into 16 standard categories, with an “Other” category for needs that do not fit any of them [18].

The French version of the COSI was recently validated in a single-centre cohort of 100 patients [20], with satisfactory linguistic and psychometric properties, including test–retest reliability. This study established the formal properties of the instrument but left three questions open. First, it is unclear whether patient-reported benefit reflects measured speech perception or captures a different dimension of rehabilitation. Second, it is unknown whether the measurement properties of the French COSI hold in a large, unselected population seen in routine practice. Third, it is unclear whether the predefined COSI categories match the listening needs that patients express. Answering these questions is necessary before PROMs can be used in routine care as a complement to audiometric testing.

The present study addressed these questions in a nationwide sample of 24,570 adults fitted with HAs. The primary aim was to determine whether patient-reported benefit, measured with the French COSI, is associated with or dissociated from speech perception in quiet and in noise, and whether it varies with patient characteristics. The second aim was to examine the measurement properties of the French COSI in routine practice (score distribution, agreement between its two subscores and, as an indicative value, internal consistency). The third aim was to measure how completely the predefined COSI categories capture the listening needs that patients spontaneously express. We hypothesised that perceived benefit would be only weakly related to speech perception, and that it would vary little with age even though older patients have poorer speech perception, indicating that the COSI captures a dimension of rehabilitation not reflected by audiometric testing. We also expected a substantial proportion of needs to fall outside the predefined categories.

## Materials and methods

### Study design and ethics

This nationwide, retrospective, cross-sectional study used routinely collected clinical data from patients seen between 1 January and 30 November 2024 in all 780 hearing-care centres of one national network (Amplifon, France), located across metropolitan France. These centres provide hearing screening, hearing assessment and HA fitting. Data were recorded by hearing-care professionals in the electronic patient file as part of routine care.

The extracted data included age, sex as recorded in the clinical file, HA experience (first-time or experienced user), speech perception test results (SIQ and SIN, before and after fitting) and COSI responses. The database was pseudonymised before transfer to Institut Pasteur. The national data protection agency (Commission Nationale de l’Informatique et des Libertés, CNIL) approved the data processing (approval number HGT/MLR/AR2410416). The study followed the REporting of studies Conducted using Observational Routinely-collected health Data (RECORD) statement [21].

### Study population and data selection

The extraction included 39,642 COSI questionnaires from 25,098 patients. We excluded questionnaires from patients aged under 18 or over 99 years (391 questionnaires, 220 patients) and with at least one need classified as screening, because they had not been fitted with HAs (480 questionnaires, 308 patients). Of the 38,771 remaining questionnaires (24,570 patients), 8,952 patients had more than one questionnaire. All intervals between successive questionnaires were shorter than one year (median 72 days), consistent with follow-up visits of the same fitting. To obtain independent observations, we kept the earliest eligible questionnaire of each patient. The final sample included 24,570 patients (Figure 1). Implausible speech-test values (20 questionnaires) were set to missing and the questionnaires were kept (see Section Missing data). Patients with missing age (n = 77) were kept and excluded only from age-related analyses.

**Figure 1.**
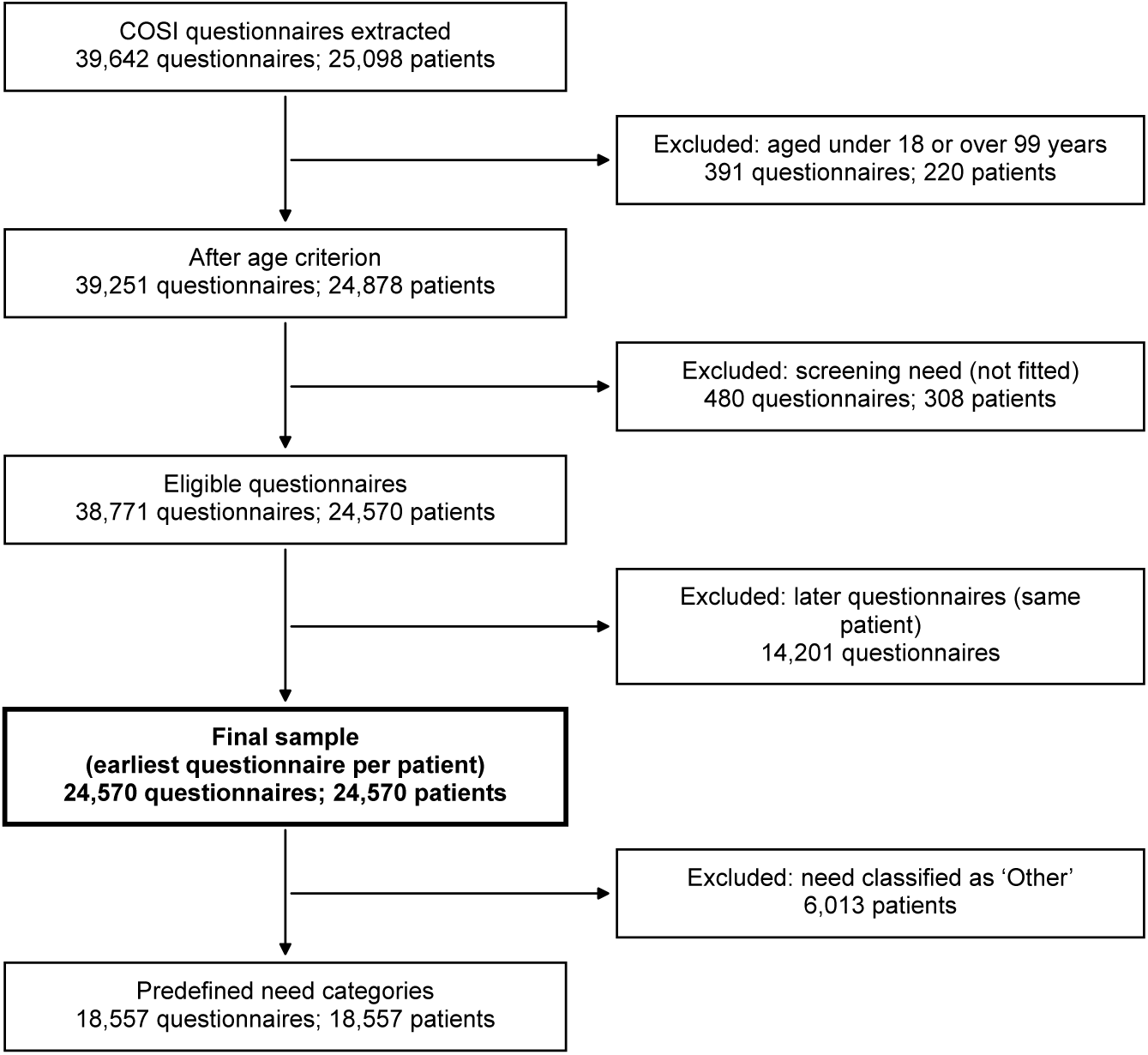
Flow chart of questionnaire and patient selection. Age exclusions were applied to each questionnaire; patients with a screening need were excluded entirely. The earliest eligible questionnaire of each patient was then kept. Implausible speech-test values were set to missing and the questionnaires were kept.

### Measures

#### Speech perception

SIQ and SIN determined the speech reception threshold (SRT): SRT in quiet is defined as the intensity level at which 50% of presented words were correctly repeated, expressed in dB HL [22]. SRT in noise is defined as the signal-to-noise ratio at which 50% of speech was correctly identified against constant background noise, expressed in dB signal-to-noise ratio (dB SNR) [23]. Higher SRT values indicate poorer performance. Values outside the plausible range (SIQ: 0 to 120 dB HL; SIN: *−*10 to +20 dB SNR) were considered data-entry errors and set to missing; the questionnaire was kept. Measured change was computed as pre-fitting minus post-fitting threshold, so that positive values indicate improvement.

Speech testing (SIQ and SIN) was not systematically performed at both time points across all centres; analyses involving SIQ or SIN were therefore restricted to patients with available data, with sample sizes reported for each analysis.

#### The French COSI

The COSI is an open, patient-driven outcome measure administered in two parts. At the initial consultation, patients identified up to five everyday specific listening needs in which they wished to improve their hearing, described in their own words and ranked by order of importance from the most important (Q1) to the least important (Q5). Following a HA trial of up to four months, patients rated, for each need, the Degree of Improvement (DI) and the Final Ability (FA) achieved with HAs, each on a five-point scale. For analysis, ordinal responses were recoded from *−*1 to 3 (Much Better/Almost Always = 3; Better/Most of the Time = 2; Slightly Better/Half the Time = 1; No Difference/Occasionally = 0; Worse/Hardly Ever = –1). The first-ranked need (Q1) was used for all analyses, except internal consistency, which required the first three ranked needs (Q1–Q3).

#### Classification of listening needs

Because the first part of the COSI is open-ended, free-text responses were mapped onto the 16 standardised listening categories defined by Dillon and colleagues. Classification was performed using CamemBERT, a transformer-based language model designed for French-language processing and built on the BERT architecture, selected for its capacity to capture contextual nuances in French-language clinical text [24]. The classification model was developed and validated on a separate, independent dataset. In that dataset, a subset of approximately 7,200 responses was manually labelled by an expert audiologist (P.M.) according to the 16 predefined categories and used to train the model; model performance was then evaluated by cross-validation and expert review of a random sample of approximately 21,600 responses, yielding a classification accuracy of 93%. On an independent hand-labelled sample (n = 1,350 responses, of which 135 were labelled “Other”), the classifier detected the “Other” category with a precision of 0.83 (95% CI 0.76 to 0.88), a recall of 0.94 (95% CI 0.89 to 0.97) and an F1 score of 0.88 (95% CI 0.84 to 0.92). Confidence intervals for precision and recall are Wilson intervals; the interval for F1 was obtained by bootstrap (2,000 resamples). The trained and validated model was subsequently applied to the present dataset to classify the COSI responses of the 39,642 extracted questionnaires (25,098 patients). In practice, the model assigned nine of the 16 original COSI categories (eight pre-defined categories and “Other”). It also assigned one additional label, “Screening”, which is not part of the original COSI taxonomy and identified patients seen for hearing screening only. The remaining categories (for example hearing traffic, the doorbell, the telephone ringing or a familiar speaker on the telephone) occurred too rarely in the labelled data to be reliably assigned and were absorbed into neighbouring categories. Analysis code is publicly available (https://github.com/mcampi111/COSI).

#### Thematic analysis of “Other” responses

First-ranked (Q1) responses assigned the residual “Other” label were further examined to assess whether this category was thematically structured rather than residual noise. We applied a rule-based thematic tagging to these responses. The coding scheme was developed from an initial reading of “Other” responses, from which recurring needs were grouped into themes. Each theme was operationalised as a set of lexical patterns (case-insensitive, accent-insensitive regular expressions applied to the normalised French text, with Unicode accent folding and light morphological normalisation). Every response was matched independently against all themes, so a single response could receive multiple tags; theme prevalence was defined as the proportion of “Other” responses matching at least one pattern of that theme. Because the scheme is non-exclusive, prevalences do not sum to 100%. The complete lexica are listed in the Supplementary Information (SI), and the tagging script is available in the public repository (https://github.com/mcampi111/COSI Validation).

### Statistical analysis

#### General approach

Categorical variables are reported as counts and percentages, and continuous variables as means and standard deviations (SD). Ordinal COSI subscores were treated as interval data for parametric tests. They were not log-transformed, because they are bounded ordinal ratings with few response categories: a monotonic transformation cannot remove a ceiling effect, as all patients at the highest score keep the same value. Each parametric test was checked with a non-parametric test (Spearman’s rho for Pearson correlations, the Wilcoxon signed-rank test for the paired t-test, the Mann–Whitney U test for Welch’s t-tests and the Kruskal–Wallis test for one-way ANOVA).

Given the large sample size, even trivial differences could reach statistical significance. Effect sizes, not p-values, were therefore the primary basis for interpreting clinical relevance; p-values are reported for completeness, with statistical significance set at two-sided p *<* 0.05. Effect sizes were interpreted with conventional thresholds [25]: Cohen’s d and dz (0.2 small, 0.5 medium, 0.8 large), *η*^2^ (0.01, 0.06, 0.14), Cramér’s V (0.1, 0.3, 0.5) and Pearson’s r (0.1, 0.3, 0.5). Main estimates (correlations, ICC, mean bias, Cronbach’s *α* and pairwise differences) are reported with their 95% confidence intervals, which were used to judge the precision of each estimate.

Pairwise comparisons after ANOVA used Tukey’s honestly significant difference test. For the *χ*^2^ test, adjusted standardised residuals were corrected for multiple testing with the Benjamini–Hochberg false discovery rate procedure. All analyses were performed in Python (version 3.9.7).

#### Missing data

Analyses were conducted on available data, without imputation or inverse probability weighting. No patient was excluded because of missing values: each analysis included all patients with data for the variables involved (available-case analysis), and the sample size is reported for each analysis. Missing data were rare for demographic variables and COSI scores (age 0.3%, sex *<*0.1%, HA experience 1.4%, DI 0.9%, FA 10.7%). They were frequent for speech tests (post-fitting SIQ 65.1%, SIN 70.3%; pre-fitting SIQ 94.2%, SIN 86.5%).

Inverse probability weighting was not applied because patients with and without a post-fitting speech test were already balanced on all available characteristics (all *|*SMD*| <* 0.1 considered negligible [26]); Table S1); weights based on these characteristics would therefore have been close to 1 and would not have changed the estimates. Multiple imputation was not used for speech tests because up to 94% of values were missing and their main determinant (centre practice) was not recorded.

#### Perceived benefit and speech perception

Pearson correlations were computed between perceived benefit (DI and FA) and postfitting SIQ and SIN, and between age and each of these four measures. Each correlation is reported with its 95% confidence interval (Fisher z transformation), Spearman’s rho as a check, and its sample size, which varied with the availability of speech tests. Higher SIQ and SIN values indicate poorer speech perception; a negative correlation therefore means that better speech perception was associated with more perceived benefit. The full correlation matrix, including pre-fitting scores, is given in the SI (Figure S1).

Because the DI rates a perceived change, it was also correlated with the measured change in speech perception (pre-fitting minus post-fitting threshold, in dB; positive values indicate improvement) in patients with both pre-and post-fitting tests.

To show how speech perception and perceived benefit vary with age, mean post-fitting SIQ and SIN, DI and FA were plotted by age group (18–50, 51–65, 66–80 and 81–99 years).

Sensitivity analyses tested whether the correlations depended on the choice of questionnaire or on the time since the first COSI interview. The correlations were repeated (1) with the last instead of the earliest eligible questionnaire of each patient, and (2) after excluding questionnaires completed less than 30 days after the first COSI interview. The date of the first COSI interview was available for 18.8% of patients only. This subgroup was therefore also analysed on its own, (3) as a whole and (4) restricted to questionnaires completed 30 days or more after the first interview, so that (4) could be compared with (3).

#### Perceived benefit across patient characteristics

DI and FA were compared between sexes and between first-time and experienced HA users with Welch’s t-test, with Cohen’s d as the effect size and the Mann–Whitney U test as a check. They were compared across four age groups (18–50, 51–65, 66–80 and 81–99 years) with one-way ANOVA, with *η*^2^ as the effect size, Tukey’s HSD test for pairwise comparisons and the Kruskal–Wallis test as a check.

#### Measurement properties

##### Ceiling effect

The distributions of DI and FA were described by the frequency of each response category and by skewness. A ceiling effect was considered present when more than 15% of patients gave the highest score [27].

##### Agreement between DI and FA

Agreement between the two subscores was assessed at three levels in patients with both scores. The mean difference was tested with a paired t-test, with Cohen’s dz as the effect size and the Wilcoxon signed-rank test as a check. Shared variance was assessed with the Pearson correlation. Absolute agreement was assessed with the intraclass correlation coefficient (two-way random effects, absolute agreement, single measurement; ICC(2,1)) and its 95% confidence interval, interpreted as poor (*<*0.50), moderate (0.50–0.75), good (0.75–0.90) or excellent (*>*0.90). A Bland–Altman plot showed the mean bias, with its 95% confidence interval, and the 95% limits of agreement.

##### Internal consistency

Each patient chooses their own listening needs, so the COSI items differ between patients. Cronbach’s *α* assumes a fixed set of items, which the COSI does not have; we therefore report it as an indicative value only. Test–retest reliability, which is better suited to an individualised instrument, could not be assessed in this retrospective design. Cronbach’s *α* was computed for DI and FA across the first three ranked needs (Q1 to Q3), in patients with all three needs completed, with its 95% confidence interval (Feldt method). It was computed overall and by sex, age group and HA experience (subgroups with at least 30 patients). Subgroups were compared through the overlap of their confidence intervals, because a smaller range of scores within a subgroup can lower *α* without any change in reliability.

##### Coverage of the COSI taxonomy

The proportion of first-ranked needs classified as “Other” was computed in the final sample. It was also corrected for classifier performance (corrected proportion = raw proportion *×* precision / recall).

Comparisons between need categories used the predefined categories only (n = 18,557), because “Other” groups heterogeneous needs. DI, FA and age were compared across categories with one-way ANOVA, with *η*^2^ as the effect size, Tukey’s HSD test for pairwise comparisons and the Kruskal–Wallis test as a check. The association between sex and need category was tested with the *χ*^2^ test, with Cramér’s V as the effect size. Adjusted standardised (Haberman) residuals identified over-and under-represented cells; their p-values were corrected with the Benjamini–Hochberg false discovery rate procedure. The same *χ*^2^ test including “Other” is reported in the SI.

## Results

### Study population

The selection of patients is shown in Figure 1, and their characteristics in Table 1. The final sample included 24,570 patients. Mean age was 75.9 years (SD = 11.9; n = 24,493). The sample comprised 12,975 women (52.8%) and 11,594 men (47.2%). First-time HA users represented 62.2% of the sample, and experienced users 37.8% (n = 24,217). Mean DI was 2.25 (SD = 0.73; n = 24,343) and mean FA was 2.19 (SD = 0.74; n = 21,938).

**Table 1.** Characteristics of the sample (one questionnaire per patient).

| Variable | Patients (n = 24,570) | Missing, n (%) |
|---|---|---|
| Age, years, mean (SD) | 75.9 (11.9) | 77 (0.3) |
| 18–50 years, n (%) | 787 (3.2) |  |
| 51–65 years, n (%) | 3,101 (12.7) |  |
| 66–80 years, n (%) | 11,386 (46.5) |  |
| 81–99 years, n (%) | 9,219 (37.6) |  |
| Sex |  | 1 (<0.1) |
| Women, n (%) | 12,975 (52.8) |  |
| Men, n (%) | 11,594 (47.2) |  |
| Hearing aid experience |  | 353 (1.4) |
| First-time users, n (%) | 15,069 (62.2) |  |
| Experienced users, n (%) | 9,148 (37.8) |  |
| Degree of Improvement (−1 to 3), mean (SD) | 2.25 (0.73) | 227 (0.9) |
| Final Ability (−1 to 3), mean (SD) | 2.19 (0.74) | 2,632 (10.7) |
| SIQ pre-fitting, dB HL, mean (SD) | 46.0 (11.9) | 23,135 (94.2) |
| SIQ post-fitting, dB HL, mean (SD) | 39.3 (8.4) | 16,007 (65.1) |
| SIN pre-fitting, dB SNR, mean (SD) | 4.4 (3.7) | 21,246 (86.5) |
| SIN post-fitting, dB SNR, mean (SD) | 2.7 (3.1) | 17,279 (70.3) |

Percentages are calculated on patients with available data. SD: standard deviation; SIQ: speech in quiet; SIN: speech in noise. Higher SIQ and SIN values indicate poorer performance.

Speech tests were available for a subset of patients only. Before fitting, mean SIQ was 46.0 dB HL (SD = 11.9; n = 1,435) and mean SIN was 4.4 dB SNR (SD = 3.7; n = 3,324). After fitting, mean SIQ was 39.3 dB HL (SD = 8.4; n = 8,563) and mean SIN was 2.7 dB SNR (SD = 3.1; n = 7,291). Patients with at least one post-fitting speech test (n = 11,447) and patients without (n = 13,123) were similar on age, sex, HA experience, DI, FA and need category (all *|*SMD*| <* 0.1; Table S1).

### Perceived benefit and speech perception

*Post-fitting speech perception.* Perceived benefit was only weakly associated with post-fitting speech perception (Table 2). Correlations were small for the DI (SIQ: r = *−*0.11, 95% CI *−*0.13 to *−*0.09, n = 8,516; SIN: r = *−*0.12, 95% CI *−*0.14 to *−*0.10, n = 7,252) and for the FA (SIQ: r = *−*0.13, n = 7,346; SIN: r = *−*0.15, n = 6,456). Spearman correlations gave similar results. In contrast, age was moderately correlated with post-fitting SIQ and SIN (both r = 0.34), but its correlation with perceived benefit was negligible (DI: r = *−*0.05; FA: r = *−*0.06). Older patients, therefore, had poorer speech perception, whereas perceived benefit was almost the same across age groups (Figure 2). The full correlation matrix, including pre-fitting scores, is shown in Figure S1.

**Figure 2.**
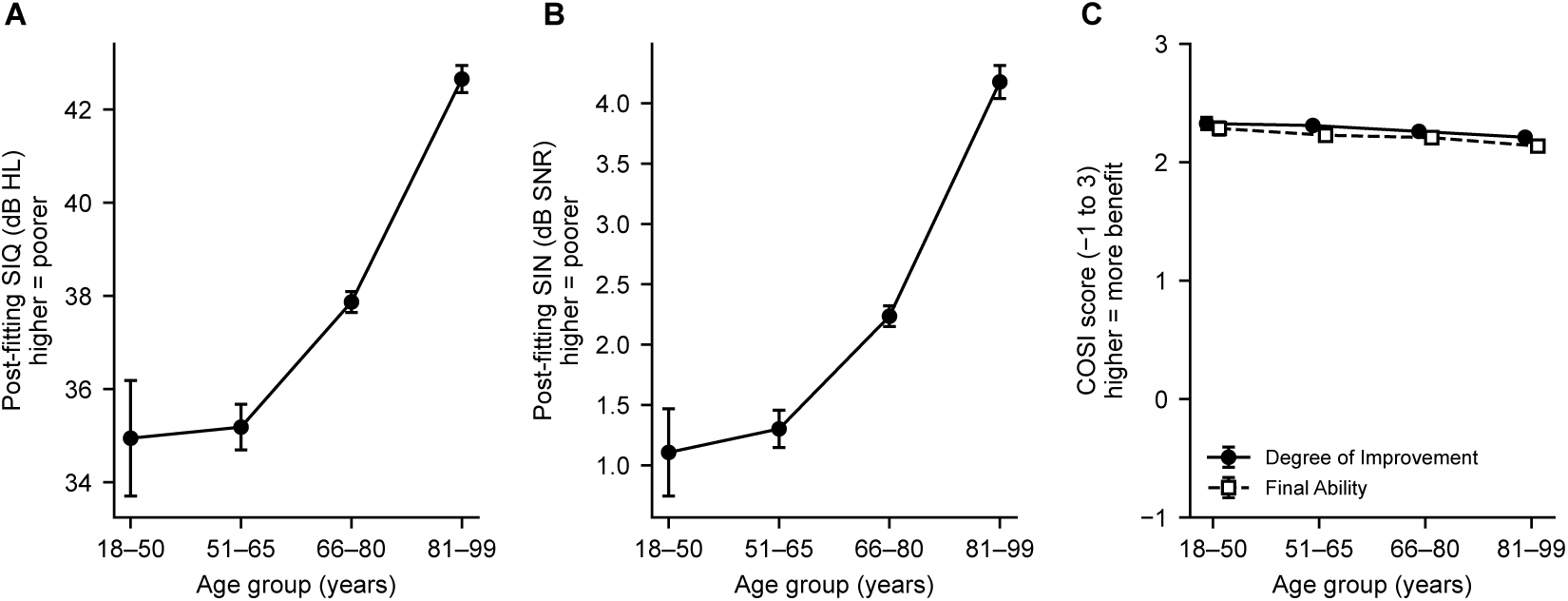
Speech perception and perceived benefit by age group. Mean (95% CI) post-fitting speech in quiet (A; n = 226, 1,020, 4,038 and 3,252 from the youngest to the oldest group), post-fitting speech in noise (B; n = 243, 1,053, 3,585 and 2,393) and COSI subscores (C; Degree of Improvement: n = 776, 3,065, 11,288 and 9,138; Final Ability: n = 707, 2,798, 10,167 and 8,195). Higher SIQ and SIN values indicate poorer speech perception. Panels A and B show only the range of observed means; panel C shows the full COSI scale.

**Table 2.**
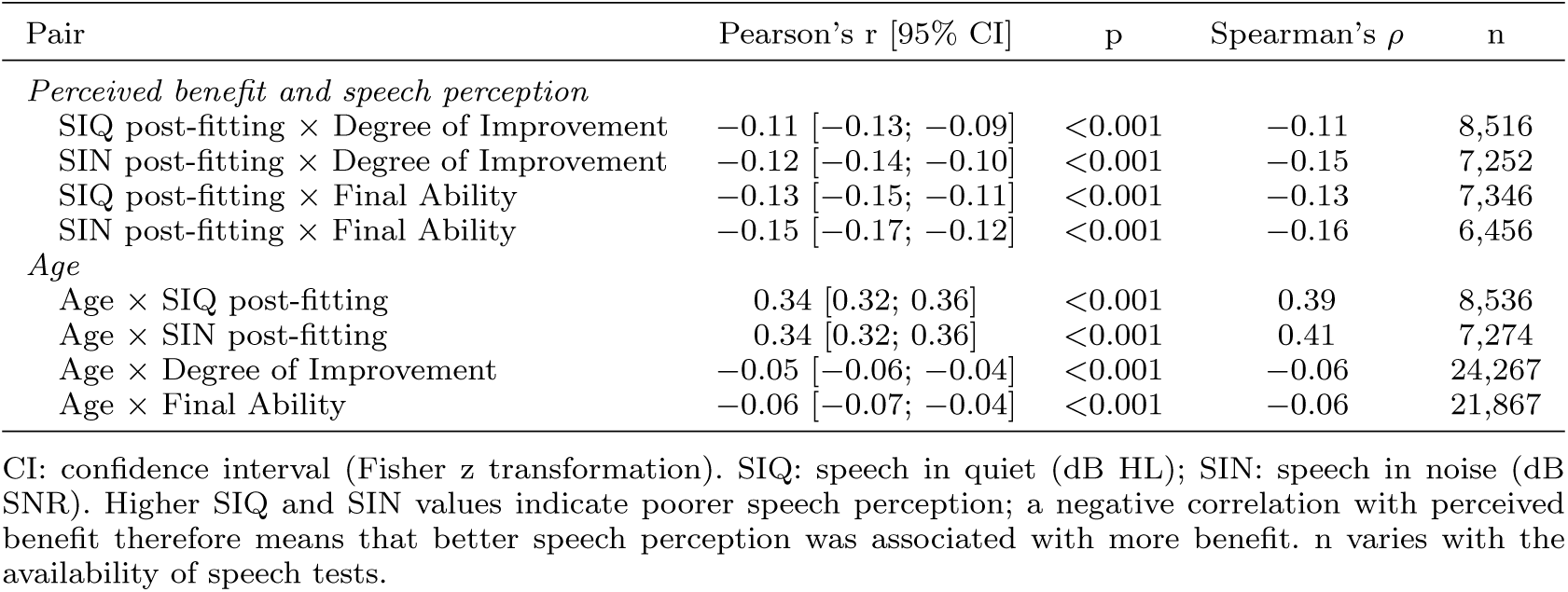
Correlations between perceived benefit, age and post-fitting speech perception.

#### Measured versus perceived change

Measured speech perception improved after fitting in most patients with both pre-and post-fitting tests (SIQ: mean change +9.1 dB, n = 614, 90% improved; SIN: +2.7 dB, n = 1,653, 81% improved). However, the measured change was not correlated with the DI in quiet (SIQ: r = 0.06, 95% CI *−*0.02 to 0.14, p = 0.14, n = 612) and only negligibly in noise (SIN: r = 0.08, 95% CI 0.03 to 0.13, p *<* 0.001, n = 1,648). Sample sizes were slightly smaller than for the mean change (614 and 1,653) because the DI was missing for some patients.

#### Sensitivity analyses

The correlations with post-fitting SIQ and SIN were almost unchanged when the last instead of the earliest questionnaire was used (for example, SIQ *×* DI: r = *−*0.13 vs *−*0.11) and after removing questionnaires completed less than 30 days after the first COSI interview (r = *−*0.11) (Table S2). The date of this interview was known for 18.8% of patients only. In this subgroup, correlations were weaker (SIQ *×* DI: r = *−*0.04) whatever the delay (r = *−*0.02 with a delay of 30 days or more), so the time since the first interview did not explain the results.

### Perceived benefit across patient characteristics

Perceived benefit did not differ between women and men (DI: 2.25 vs 2.25, Welch’s t(24,037) = 0.13, p = 0.90, d = 0.00; FA: 2.18 vs 2.19, t(21,678) = *−*0.34, p = 0.73, d = *−*0.01), nor between first-time and experienced users (DI: 2.25 vs 2.26, t(19,140) = *−*0.24, p = 0.81, d = 0.00; FA: 2.18 vs 2.20, t(17,330) = *−*1.17, p = 0.24, d = *−*0.02). Mann–Whitney tests gave the same results.

Perceived benefit was slightly lower in older age groups, from 2.33 (18–50 years) to 2.21 (81–99 years) for DI and from 2.29 to 2.14 for FA. These differences were statistically significant but negligible (DI: F(3, 24,263) = 19.53, p *<* 0.001, *η*^2^ = 0.002; FA: F(3, 21,863) = 22.80, p *<* 0.001, *η*^2^ = 0.003), and the Kruskal–Wallis tests gave the same results. In pairwise comparisons, the 81–99 group scored lower than all other groups, by 0.05 to 0.12 points for DI and 0.07 to 0.15 points for FA (Table S3).

### Measurement properties

#### Ceiling effect

Both subscores showed a ceiling effect. The highest score was given by 39.9% of patients for DI (n = 24,343) and 35.4% for FA (n = 21,938), more than twice the 15% criterion. Overall, 87.4% of patients rated their improvement as “better” or “much better”. Both distributions were negatively skewed (skewness *−*0.77 and *−*0.86).

#### Agreement between DI and FA

Among the 21,711 patients with both subscores, mean DI was higher than mean FA (2.31 vs 2.19; mean difference 0.12; paired t(21,710) = 35.43, p *<* 0.001; Wilcoxon p *<* 0.001). This difference was small (Cohen’s dz = 0.24). The two subscores shared about half of their variance (r = 0.73; r^2^ = 0.54). Absolute agreement was moderate (ICC(2,1) = 0.72, 95% CI 0.69 to 0.75). The Bland–Altman analysis showed a mean bias of 0.12 (95% CI 0.12 to 0.13) and 95% limits of agreement from *−*0.89 to 1.13, a range of about two points on the five-point scale (Figure S2). The two subscores were identical for 75.6% of patients and differed by one point for 23.3% (DI higher in 17.8%, FA higher in 5.5%) (Table S4).

#### Internal consistency

Across the first three ranked needs, Cronbach’s *α* was 0.808 (95% CI 0.801 to 0.815; n = 9,126) for DI and 0.806 (95% CI 0.799 to 0.813; n = 8,095) for FA. Values were similar across sex, age groups and HA experience (range 0.776 to 0.825), with overlapping confidence intervals (Table S5).

### Coverage of the COSI taxonomy

#### Distribution of needs

The model assigned the first-ranked needs to eight predefined categories and to “Other”. The most frequent predefined categories were conversation with one or two people in quiet (21.4%), conversation with a group in noise (21.3%) and television or radio at normal volume (12.8%), followed by conversation with a group in quiet (8.4%) and feeling embarrassed or stupid (7.0%) (Figure 3A; Table S6). “Other” accounted for 24.5% of first-ranked needs (n = 6,013), or 21.6% after correction for classifier performance.

**Figure 3.**
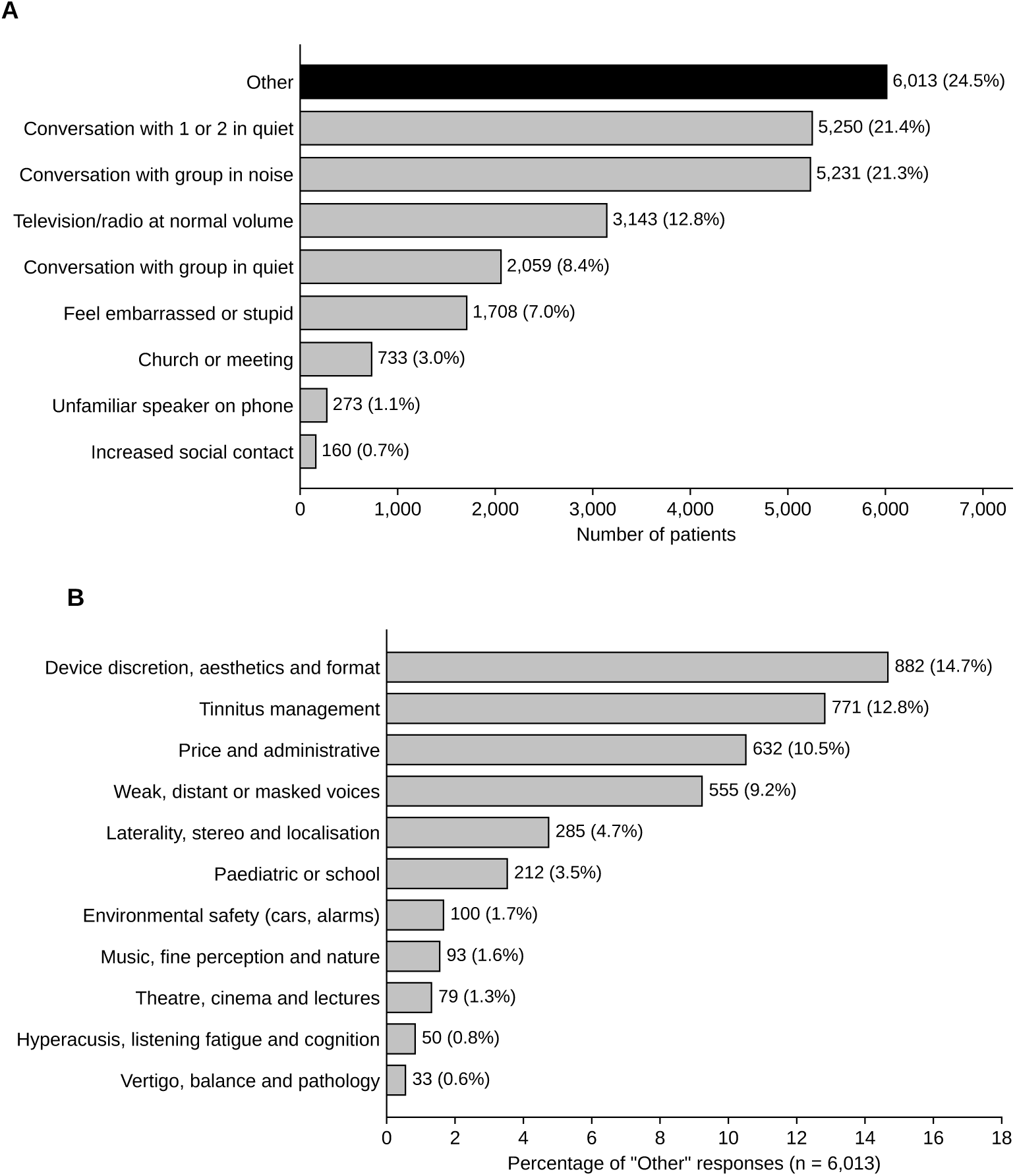
Distribution of the first-ranked COSI needs (n = 24,570 patients). (A) Number and percentage of patients in each need category; “Other” (black) groups the needs that did not match any predefined category. (B) Prevalence of the themes identified within the first-ranked “Other” responses (n = 6,013); a response could receive several themes, so percentages do not sum to 100.

#### Themes within “Other”

Rule-based thematic tagging of the “Other” responses identified eleven recurring themes. Among the 6,013 first-ranked “Other” responses, the most prevalent themes were device discretion and design (14.7%; e.g. “hearing aids should be invisible”), tinnitus (12.8%; e.g. “reduce my tinnitus”), price and administrative aspects (10.5%), weak or distant voices (9.2%; e.g. “hearing someone across the room”) and sound localisation (4.7%) (Figure 3B; the lexicon of each theme is given in Table S9). Because tagging was non-exclusive, prevalences do not sum to 100%.

#### Comparisons between predefined categories (n = 18,557)

Mean age differed across categories, from 67.9 years (church or meeting) to 78.4 years (unfamiliar speaker on the telephone) (F(7, 18,491) = 88.04, p *<* 0.001, *η*^2^ = 0.03; Kruskal–Wallis H = 538.2, p *<* 0.001). Patients whose first need was church or meeting were 7 to 11 years younger than patients in every other category. Perceived benefit also differed, but the effects were negligible (DI: from 2.10 to 2.34, F(7, 18,392) = 17.16, p *<* 0.001, *η*^2^ = 0.006; FA: from 2.09 to 2.26, F(7, 16,587) = 12.07, p *<* 0.001, *η*^2^ = 0.005) (Table S7). The lowest DI was reported for the telephone with an unfamiliar speaker (2.10). Sex was associated with need category, but the association was weak (*χ*^2^(7) = 151.9, p *<* 0.001, Cramér’s V = 0.09). Women more often reported the telephone with an unfamiliar speaker (73.3% women; adjusted residual +7.0), feeling embarrassed or stupid (+6.1), television or radio (+4.1) and conversation with a group in quiet (+3.4), and men more often reported conversation with a group in noise (+6.4) and conversation with one or two people in quiet (+4.6); all these residuals remained significant after Benjamini–Hochberg correction (Table S8). Including “Other” gave similar results (*χ*^2^(8) = 157.9, V = 0.08; Table S8).

## Discussion

In this nationwide sample of 24,570 adults fitted with HAs in 780 centres, perceived benefit measured with the French COSI was only weakly associated with speech perception. Correlations between perceived benefit and post-fitting speech perception were small (r = *−*0.11 to *−*0.15), and perceived change was not or only negligibly related to measured change in speech perception (r = 0.06 in quiet and 0.08 in noise). Older patients had poorer speech perception (r = 0.34), whereas perceived benefit varied very little with age, sex or HA experience (all effect sizes negligible). Benefit scores were high (87.4% of patients rated their improvement as “better” or “much better”) and showed a clear ceiling effect, which limits the variance available for these associations. The two COSI subscores agreed on average but were not interchangeable for all patients (ICC = 0.72), and internal consistency, reported as an indicative value, was 0.81 for both subscores. Finally, about one patient in four (24.5%; 21.6% after correction for classifier performance) expressed a first-ranked need that did not fit any of the predefined COSI categories. These results confirm both hypotheses: perceived benefit was largely dissociated from speech perception, and a substantial share of patients’ needs fell outside the predefined COSI categories.

### Perceived benefit and speech perception

Older patients had poorer post-fitting speech perception, in quiet and in noise, whereas age was almost unrelated to perceived benefit. Perceived benefit itself was only weakly associated with post-fitting speech perception. This dissociation is consistent with previous reports of no clear relationship between speech perception outcomes and HA satisfaction [11], and with studies reporting no association between age and HA satisfaction [28, 29].

The most direct test of whether perceived benefit follows measured benefit is the correlation between perceived change (DI) and measured change in speech perception (pre-fitting minus post-fitting threshold). In patients with both tests, measured speech perception improved in most cases (90% in quiet and 81% in noise), but this improvement was not related to perceived change in quiet (r = 0.06, n = 612) and only negligibly in noise (r = 0.08, n = 1,648). Patients who improved most on speech tests therefore did not report more benefit than the others. These results were stable in sensitivity analyses: the correlations were almost unchanged when the last instead of the earliest questionnaire was used, and after removing questionnaires completed less than 30 days after the first COSI interview (Table S2).

These findings should nevertheless be interpreted with caution. Perceived benefit was strongly skewed towards the top of the scale (skewness *−*0.77 and *−*0.86), and this ceiling effect reduces its variance. Weak correlations may therefore partly reflect this reduced variance, and the present data cannot show on their own that the COSI measures a construct fully independent of speech perception. The ceiling effect is unlikely to explain all of the dissociation, however: about 60% of patients did not give the highest score, the correlations were based on several thousand patients, and age, which was moderately related to speech perception, was almost unrelated to perceived benefit.

Within these limits, the stability of perceived benefit across age is notable. Older patients may adjust their expectations, or judge the success of their fitting on criteria less dependent on speech test scores, such as specific everyday situations. Perceived benefit may thus reflect real-world communication needs and adaptation strategies that audiometric tests do not capture, which supports the combined use of patient-reported and audiological measures [30, 31].

### Perceived benefit across patient characteristics

Perceived benefit did not differ between women and men, in agreement with previous studies [31–33]. It was slightly lower in the oldest age group, but age explained a negligible proportion of the variance (*η*^2^ *≤* 0.003; at most 0.15 points on the *−*1 to 3 scale). This is consistent with the literature reporting no clear relationship between chronological age and subjective benefit [19, 30, 32], even though objective benefit may decrease with age [33, 34].

Perceived benefit did not differ between first-time and experienced users either (d = 0.00 for the DI and *−*0.02 for the FA). This result is consistent with findings from the International Outcome Inventory for Hearing Aids, in which experienced users often report greater benefit but also more residual difficulties, resulting in comparable overall outcomes [35, 36].

### Measurement properties

Both COSI subscores showed a clear ceiling effect: the highest score was given by 39.9% of patients for the DI and 35.4% for the FA, more than twice the usual 15% criterion. This is consistent with Dillon et al.[19], who also reported benefit scores skewed towards high values. Benefit may also be overestimated when clinicians are involved in goal setting and scoring [19, 37]. A ceiling effect reduces the ability of the COSI to detect differences between patients or groups, and limits its use as an outcome measure for comparisons, which was not its original purpose [38].

The two subscores, DI and FA, were closely related but not interchangeable. Their mean difference was statistically significant because of the large sample, but small (0.12 points; Cohen’s dz = 0.24). The two subscores were identical for three patients out of four (75.6%), and differed by one point for almost all the others (23.3%), more often in favour of the DI. Absolute agreement was moderate (ICC = 0.72, 95% CI 0.69 to 0.75), and the two subscores shared about half of their variance (r = 0.73). This pattern is expected, because the FA partly depends on the improvement obtained. Lansbergen et al. [39] suggested that the FA alone may be enough to evaluate COSI goals; the present results qualify this suggestion, because the two subscores differ for about one patient in four and therefore provide partly different information.

The internal consistency of the subscores across the first three needs was 0.81. This value should be interpreted with caution: because each patient chooses their own listening situations, *α* reflects the consistency with which a patient rates their own needs rather than the reliability of a fixed set of items.

#### Comparison with the single-centre French validation

The French COSI was first validated by Tuset et al.[20] in a prospective, single-centre cohort of 100 HA users, who reported a Cronbach’s *α* of 0.76 to 0.79 across subscales, a test–retest reliability of 0.68 to 0.81 and moderate convergent validity with the IOI-HA. The indicative *α* observed here (0.81) was of the same order, in a sample about 250 times larger and drawn from routine practice. The two studies therefore provide complementary evidence: the single-centre study established the formal psychometric properties of the instrument, including the temporal stability that a retrospective design cannot assess, while the present study describes its behaviour at national scale and in real-world conditions. They are also methodologically complementary. Tuset et al. classified open-ended responses manually in a small sample, whereas the present study used automated classification with a French-language transformer model, which is necessary for the population-scale analysis of an individualised instrument.

### Coverage of the COSI taxonomy

About one patient in four expressed a first-ranked need classified as “Other” (24.5%; 21.6% after correction for classifier performance). This proportion is at least as high as the 21.2% reported in the original COSI studies [18, 19], three decades after the taxonomy was defined. The predefined categories may therefore no longer capture a substantial share of current patients’ priorities.

Thematic tagging showed that “Other” was not residual noise but contained recurring needs that are absent from the predefined categories, such as device discretion and design (14.7%), tinnitus (12.8%), cost and administrative aspects (10.5%), distant or weak voices (9.2%) and sound localisation (4.7%). Tinnitus alone concerned about one “Other” response in eight, but has no place in the current taxonomy. These findings agree with previous studies that pointed to similar limitations and to the difficulty for clinicians to classify some responses reliably [40, 41]. Updating the taxonomy, by grouping rarely used categories and adding current domains such as tinnitus, device design, cost or remote communication, could improve its relevance and content validity for current patients, while keeping the open-ended format that captures the full range of patient experience.

This thematic analysis is descriptive and exploratory. The coding scheme was developed from the reading of “Other” responses, and the lexical rules were not validated against an independent manual annotation. The prevalences should therefore be read as an approximate ordering of these needs rather than as precise frequencies.

#### Listening needs by age and sex

The distribution of first-ranked needs varied with age. Patients whose first need was to understand at church or meeting were the youngest, 7 to 11 years younger than patients in every other category, whereas those reporting the telephone with unfamiliar speakers were the oldest. Younger patients may thus be more concerned with social and professional participation, and older patients with everyday functional communication. Qualitative work suggests that older adults may normalise hearing impairment as part of ageing and focus on functional communication [42], and the psychosocial impact of HL is known to vary with age [43]. Listening goals may therefore follow the patient’s stage of life, which supports tailored rehabilitation.

Sex was associated with need category, but the association was weak (Cramér’s V = 0.09). Women more often reported the telephone with unfamiliar speakers, feeling embarrassed or stupid, television or radio and conversation with a group in quiet, whereas men more often reported conversation with a group in noise and conversation with one or two people in quiet. These patterns may reflect differences in communication habits or social roles and would need dedicated investigation.

Perceived benefit differed little across need categories (*η*^2^ *<* 0.01). The lowest DI was reported for the telephone with unfamiliar speakers (2.10, vs 2.34 for group conversation in quiet). The telephone remains one of the most difficult situations for HA users, because of restricted bandwidth, the lack of binaural cues and signal distortion [44]. Targeted counselling and connectivity solutions, such as Bluetooth streaming, may therefore deserve systematic attention during rehabilitation. This interpretation should remain cautious, because this category concerned only 273 patients and the effect sizes were negligible.

### Strengths and limitations

The main strength of this study is its size: to our knowledge, it is one of the largest evaluations of a hearing-related PROM, covering routine practice in 780 centres and a broad range of patient profiles. Keeping one questionnaire per patient ensured independent observations, and the main results were stable in sensitivity analyses. Automated classification of free-text responses made it possible to analyse an open-ended instrument at population scale, which would not be feasible manually.

Several limitations should be considered. First, benefit scores showed a strong ceiling effect, which reduces the variance available for correlations and comparisons, and may partly explain the weak associations with speech perception. Models designed for bounded outcomes, such as censored (Tobit) regression, two-part models separating patients at the ceiling from the others, or ordinal regression, could test whether the weak associations persist after accounting for the ceiling; they should be used in future work or as sensitivity analyses.

Second, speech tests were not performed systematically in all centres: data were missing for 65% to 94% of patients depending on the test. The analysed subsets remained large (1,435 to 8,563 patients), and patients with and without a post-fitting speech test were similar on age, sex, HA experience, COSI scores and need category (all *|*SMD*| <* 0.1; Table S1). They may still differ on variables that were not available in the dataset, such as the centre or the degree of HL.

Third, the comparison between perceived and measured change, which is the most direct test of the dissociation, was limited to the patients with both pre-and post-fitting tests (n = 612 in quiet and 1,648 in noise). Similarly, the date of the first COSI interview was known for 18.8% of patients only, and correlations were weaker in this subgroup. The precision and generalisability of these analyses are therefore limited, although their results agree with those observed at the post-fitting level.

Fourth, pure-tone thresholds were not available, so the analyses could not account for the degree of HL. Fifth, need categories were assigned by an automated classifier; classification errors are possible, although its performance for the “Other” category was high (precision 0.83, recall 0.94), and the proportion of “Other” was corrected accordingly. Sixth, the retrospective design did not allow the assessment of test–retest reliability; this property was documented in the single-centre French validation [20]. Finally, all data came from a single national hearing-care network, which may limit generalisability to other healthcare systems.

Future studies should collect speech tests and pure-tone thresholds systematically before and after fitting, so that perceived and measured benefit can be compared in all patients and adjusted for the degree of HL. A longitudinal design would also allow test–retest reliability and changes in perceived benefit over time to be assessed. Finally, an updated COSI taxonomy should be developed and tested on new data.

### Clinical implications

Because perceived benefit was only weakly related to speech perception, the COSI and speech tests should be used together rather than one in place of the other: speech tests describe what the patient can hear, and the COSI describes whether the patient’s own goals are met. Speech test scores alone should not be used to judge the success of a fitting for an individual patient. Because the two COSI subscores differ for about one patient in four, both should be recorded. Clinicians should also pay particular attention to needs that the predefined categories do not cover, such as tinnitus, device discretion or cost, and to situations with lower benefit, such as the telephone.

## Conclusions

In routine practice, perceived benefit measured with the French COSI was high and only weakly related to speech perception or to measured improvement, and it varied very little with patient characteristics; a strong ceiling effect should be considered when interpreting these results. The two COSI subscores agreed on average but were not interchangeable for all patients. About one patient in four expressed a first-ranked need outside the predefined categories, which supports an update of the COSI taxonomy.

## Acknowledgements

The authors thank Amplifon France for assistance with data collection. Further, the authors thank the Clinical Research Coordination Office (CRCO) of the Institut Pasteur for their help with biomedical regulatory and ethical aspects of the project.

## Declaration of generative AI use

The authors report generative AI was not used in their research or preparation of this manuscript.

## Data availability statement

The data provider is Amplifon France. The data generated and analysed during the current study are available from the corresponding author upon reasonable request.

## Abbreviations

ASHA: American Speech-Language-Hearing Association;
COSI: Client Oriented Scale of Improvement;
DI: Degree of Improvement;
FA: Final Ability;
HA: hearing aid;
HL: Hearing loss;
PROMS: patient-reported outcome measures;
RECORD: REporting of studies Conducted using Observational Routinely-collected health Data;
SD: standard deviations;
SIQ: speech in quiet;
SIN: speech in noise;
SMD: standardised mean differences;
SRT: speech reception threshold;
WHO: World Health Organization;

