## Supplemental Information for "Patient-reported benefit and speech perception in 24,570 hearing aid users: a nationwide study of the French Client-Oriented Scale of Improvement (COSI)"

RESEARCH ARTICLE

[a]Université Paris Cité, Institut Pasteur, AP-HP, INSERM, CNRS, Fondation Pour l'Audition, Institut de l'Audition, IHU reConnect, Paris, F-75012, France; [b]Amplifon, Paris, France; [c]Deep Hearing Lab, University of Zurich and University Hospital Zurich, Zurich, Switzerland; [d]Department of Statistics and Applied Probability, University of California Santa Barbara, Santa Barbara, CA 93106, United States; [e]Department of Audiology and Neurotology, Civil Hospitals of Lyon, 69003 Lyon, France; [f]Department of Physiology, Claude Bernard University, 69003 Lyon, France

**ARTICLE HISTORY**

Compiled November 12, 2026

**Abbreviations:** ASHA: American Speech-Language-Hearing Association; COSI: Client Oriented Scale of Improvement; DI: Degree of Improvement; FA: Final Ability; HA: hearing aid; HL: hearing loss; PROMs: patient-reported outcome measures; RECORD: REporting of studies Conducted using Observational Routinely-collected health Data; SD: standard deviation; SIQ: speech in quiet; SIN: speech in noise; SMD: standardised mean difference; SRT: speech reception threshold; WHO: World Health Organization

CONTACT Perrine Morvan.

### Introduction

This Supplementary Information is in support of the paper **"Patient-reported benefit and speech perception in 24,570 hearing aid users: a nationwide study of the French Client-Oriented Scale of Improvement (COSI)".** We provide all the results of our analyses described in the main body of the paper.

### Materials and methods

#### *Study design and ethics*

The study followed the REporting of studies Conducted using Observational Routinely-collected health Data (RECORD) statement [1]. The completed checklist is given below.

| | Item No. | STROBE Items | Location in manuscript | RECORD Items | Location in manuscript |
|---|---|---|---|---|---|
| **Title and abstract** | | | | | |
| | 1(a) | Indicate the study's design with a commonly used term in the title or the abstract | p. 1, Abstract (Design: nationwide, retrospective, cross-sectional study) | RECORD 1.1: The type of data used should be specified in the title or abstract. When possible, the name of the databases used should be included. | p. 1, Abstract (routinely collected clinical data); data source named in Methods, p. 4 |
| | 1(b) | Provide in the abstract an informative and balanced summary of what was done and what was found | p. 1–2, Abstract | RECORD 1.2: If applicable, the geographic region and timeframe within which the study took place should be reported in the title or abstract. | p. 1, Title (nationwide) and Abstract (780 French hearing-care centres; 2024) |

*Continued on next page*

RECORD checklist – continued from previous page

| | Item No. | STROBE Items | Location in manuscript | RECORD Items | Location in manuscript |
|---|---|---|---|---|---|
| | | | | RECORD 1.3: If linkage between databases was conducted for the study, this should be clearly stated in the title or abstract. | NA (no linkage) |
| **Introduction** | | | | | |
| Background rationale | 2 | Explain the scientific background and rationale for the investigation being reported | p. 3–4, Introduction | | |
| Objectives | 3 | State specific objectives, including any prespecified hypotheses | p. 4, Introduction (three aims and hypotheses, last paragraph) | | |
| **Methods** | | | | | |
| Study Design | 4 | Present key elements of study design early in the paper | p. 4, Study design and ethics | | |
| Setting | 5 | Describe the setting, locations, and relevant dates, including periods of recruitment, exposure, follow-up, and data collection | p. 4, Study design and ethics (780 centres; 1 January to 30 November 2024) | | |
| **Participants** | | | | | |
| *Continued on next page* | | | | | |

| | Item No. | STROBE Items | Location in manuscript | RECORD Items | Location in manuscript |
|---|---|---|---|---|---|
| | 6(a) | Cross-sectional study – Give the eligibility criteria, and the sources and methods of selection of participants | p. 5, Study population and data selection; Figure 1 (p. 12) | RECORD 6.1: The methods of study population selection (such as codes or algorithms used to identify subjects) should be listed in detail. If this is not possible, an explanation should be provided.<br><br>RECORD 6.2: Any validation studies of the codes or algorithms used to select the population should be referenced. If validation was conducted for this study and not published elsewhere, detailed methods and results should be provided. | p. 5, Study population and data selection (age criterion, exclusion of screening, earliest questionnaire per patient); p. 6–7, Classification of listening needs (“Screening” label)<br><br>p. 6–7, Classification of listening needs (development and validation of the classifier; precision, recall and F1 with 95% CI) |

*Continued on next page*

| | **Item No.** | **STROBE Items** | **Location in manuscript** | **RECORD Items** | **Location in manuscript** |
|---|---|---|---|---|---|
| | | | | RECORD 6.3: If the study involved linkage of databases, consider use of a flow diagram or other graphical display to demonstrate the data linkage process, including the number of individuals with linked data at each stage. | NA (no linkage) |
| | 6(b) | Cohort / case-control study – For matched studies, give matching criteria | NA (cross-sectional, no matching) | | |
| **Variables** | | | | | |
| Variables | 7 | Clearly define all outcomes, exposures, predictors, potential confounders, and effect modifiers. Give diagnostic criteria, if applicable. | p. 4–7, Study design and ethics (extracted variables), Measures (speech perception, the French COSI, classification of listening needs) | RECORD 7.1: A complete list of codes and algorithms used to classify exposures, outcomes, confounders, and effect modifiers should be provided. If these cannot be reported, an explanation should be provided. | p. 6–7, Classification of listening needs and Thematic analysis of "Other" responses; SI, Table S5 (categories and French labels) and Table S8 (lexica); public code repository (p. 7) |
| **Data sources/measurement** | | | | | |

*Continued on next page*

| | Item No. | STROBE Items | Location in manuscript | RECORD Items | Location in manuscript |
|---|---|---|---|---|---|
| Data sources/ measurement | 8 | For each variable of interest, give sources of data and details of methods of assessment (measurement). Describe comparability of assessment methods if there is more than one group | p. 4–7, Study design and ethics; Measures (speech perception, the French COSI, classification of listening needs) | | |
| **Bias** | | | | | |
| Bias | 9 | Describe any efforts to address potential sources of bias | p. 5 (one questionnaire per patient); p. 8–9, Missing data (comparison of patients with and without speech tests, SI Table S1); p. 9, sensitivity analyses; p. 11 (correction of “Other” for classifier performance); p. 22–23, Strengths and limitations | | |
| **Study size** | | | | | |

*Continued on next page*

| | Item No. | STROBE Items | Location in manuscript | RECORD Items | Location in manuscript |
|---|---|---|---|---|---|
| Study size | 10 | Explain how the study size was arrived at | p. 5, Study population and data selection (all eligible patients of the network); Figure 1 (p. 12) | | |
| **Quantitative variables** | | | | | |
| Quantitative variables | 11 | Explain how quantitative variables were handled in the analyses. If applicable, describe which groupings were chosen, and why | p. 5–6 (speech tests; COSI scores recoded from −1 to 3); p. 8, General approach (ordinal scores treated as interval data, no transformation); p. 9–10 (age groups) | | |
| **Statistical methods** | | | | | |
| Statistical methods | 12(a) | Describe all statistical methods, including those used to control for confounding | p. 8–11, Statistical analysis | | |

*Continued on next page*

| | Item No. | STROBE Items | Location in manuscript | RECORD Items | Location in manuscript |
|---|---|---|---|---|---|
| | 12(b) | Describe any methods used to examine subgroups and interactions | p. 10, Perceived benefit across patient characteristics and Internal consistency (subgroups); p. 11, Coverage of the COSI taxonomy | | |
| | 12(c) | Explain how missing data were addressed | p. 8–9, Missing data; SI, Table S1 | | |
| | 12(d) | Cross-sectional study – If applicable, describe analytical methods taking account of sampling strategy | p. 4–5 (all centres of the network; earliest questionnaire per patient); no sampling weights needed | | |
| | 12(e) | Describe any sensitivity analyses | p. 9, Perceived benefit and speech perception (sensitivity analyses); SI, Table S2 | | |
| **Data access and cleaning methods** | | | | | |

*Continued on next page*

| | **Item No.** | **STROBE Items** | **Location in manuscript** | **RECORD Items** | **Location in manuscript** |
|---|---|---|---|---|---|
| Data access and cleaning methods | | | | RECORD 12.1: Authors should describe the extent to which the investigators had access to the database population used to create the study population.<br>RECORD 12.2: Authors should provide information on the data cleaning methods used in the study. | p. 4–5, Study design and ethics (pseudonymised extraction transferred to Institut Pasteur; CNIL approval)<br>p. 5, Study population and data selection (age range, screening, repeated questionnaires, implausible speech-test values set to missing); Figure 1 (p. 12) |
| **Linkage** | | | | | |

*Continued on next page*

RECORD checklist – continued from previous page

| | Item No. | STROBE Items | Location in manuscript | RECORD Items | Location in manuscript |
|---|---|---|---|---|---|
| Linkage | | | | RECORD 12.3: State whether the study included person-level, institutional-level, or other data linkage across two or more databases. The methods of linkage and methods of linkage quality evaluation should be provided. | NA (single database, no linkage) |
| **Results** | | | | | |
| *Participants* | | | | | |
| Participants | 13(a) | Report the numbers of individuals at each stage of the study (e.g., numbers potentially eligible, examined for eligibility, confirmed eligible, included in the study, completing follow-up, and analysed) | p. 5 and p. 11, Study population; Figure 1 (p. 12) | | |
| | 13(b) | Give reasons for non-participation at each stage | p. 5; Figure 1 (p. 12) | | |

*Continued on next page*

| | **Item No.** | **STROBE Items** | **Location in manuscript** | **RECORD Items** | **Location in manuscript** |
|---|---|---|---|---|---|
| | 13(c) | Consider use of a flow diagram | Figure 1 (p. 12) | RECORD 13.1: Describe in detail the selection of the persons included in the study (i.e., study population selection) including filtering based on data quality, data availability and linkage. The selection of included persons can be described in the text and/or by means of the study flow diagram. | p. 5, Study population and data selection; Figure 1 (p. 12) |
| *Descriptive data* | | | | | |
| Descriptive data | 14(a) | Give characteristics of study participants (e.g., demographic, clinical, social) and information on exposures and potential confounders | p. 11–12, Study population; Table 1 | | |
| | 14(b) | Indicate the number of participants with missing data for each variable of interest | Table 1 (p. 12, column “Missing”); p. 8, Missing data | | |

*Continued on next page*

| | Item No. | STROBE Items | Location in manuscript | RECORD Items | Location in manuscript |
|---|---|---|---|---|---|
| | 14(c) | Cohort study – summarise follow-up time | NA (cross-sectional) | | |
| *Outcome data* | | | | | |
| Outcome data | 15 | Cross-sectional study – Report numbers of outcome events or summary measures | p. 11–17, Results; Table 1, Table 2, Figures 2 and 3 | | |
| *Main results* | | | | | |
| Main results | 16(a) | Give unadjusted estimates and, if applicable, confounder-adjusted estimates and their precision (e.g., 95% confidence interval). Make clear which confounders were adjusted for and why they were included | p. 13–17, Results; Table 2 (unadjusted estimates with 95% CI; no confounder adjustment, as the aims were descriptive and correlational) | | |
| | 16(b) | Report category boundaries when continuous variables were categorized | p. 9–10 (age groups 18–50, 51–65, 66–80 and 81–99 years); Table 1 | | |

*Continued on next page*

| | **Item No.** | **STROBE Items** | **Location in manuscript** | **RECORD Items** | **Location in manuscript** |
|---|---|---|---|---|---|
| | 16(c) | If relevant, consider translating estimates of relative risk into absolute risk for a meaningful time period | NA | | |
| *Other analyses* | | | | | |
| Other analyses | 17 | Report other analyses done – e.g., analyses of subgroups and interactions, and sensitivity analyses | p. 13 (sensitivity analyses); p. 14–17 (subgroups); SI, Tables S2, S3, S6 and S7 | | |
| **Discussion** | | | | | |
| Key results | 18 | Summarise key results with reference to study objectives | p. 17–18, Discussion (first paragraph) | | |

*Continued on next page*

RECORD checklist – continued from previous page

| | Item No. | STROBE Items | Location in manuscript | RECORD Items | Location in manuscript |
|---|---|---|---|---|---|
| Limitations | 19 | Discuss limitations of the study, taking into account sources of potential bias or imprecision. Discuss both direction and magnitude of any potential bias | p. 22–23, Strengths and limitations | RECORD 19.1: Discuss the implications of using data that were not created or collected to answer the specific research question(s). Include discussion of misclassification bias, unmeasured confounding, missing data, and changing eligibility over time, as they pertain to the study being reported. | p. 22–23, Strengths and limitations (ceiling effect, missing speech tests, unavailable variables such as centre and degree of hearing loss, classification errors, retrospective design, single network) |
| Interpretation | 20 | Give a cautious overall interpretation of results considering objectives, limitations, multiplicity of analyses, results from similar studies, and other relevant evidence | p. 17–22, Discussion | | |
| Generalisability | 21 | Discuss the generalisability (external validity) of the study results | p. 23, Strengths and limitations (single national hearing-care network) | | |
| **Other Information** | | | | | |
| | | | | | *Continued on next page* |

RECORD checklist – continued from previous page

| | Item No. | STROBE Items | Location in manuscript | RECORD Items | Location in manuscript |
|---|---|---|---|---|---|
| Funding | 22 | Give the source of funding and the role of the funders for the present study and, if applicable, for the original study on which the present article is based | p. 24, Funding | | |
| Accessibility of protocol, raw data, and programming code | | | | RECORD 22.1: Authors should provide information on how to access any supplemental information such as the study protocol, raw data, or programming code. | p. 7 (public code repository); p. 25, Data availability statement; SI, Table S8 (lexica) |

*Page numbers (p.) refer to the main manuscript; SI refers to the Supplementary Information. NA: not applicable.*

#### ***Statistical analysis***

*Missing data*

To assess the risk of selection bias, patients with at least one post-fitting speech test (SIQ or SIN) were compared with patients without, on age, sex, hearing aid experience, COSI scores and need category, using standardised mean differences (SMD; $|SMD| < 0.1$ considered negligible) (Table S1).

### Results

#### *Study population*

*Comparison of patients with and without a post-fitting speech test*

Speech tests were not performed systematically in all centres: a post-fitting speech test (SIQ or SIN) was available for 11,447 of the 24,570 patients (46.6%). Analyses involving speech perception were therefore based on a subset of patients. Table S1 tests whether this subset differed from the patients without a speech test, which could bias the results.

The two groups were compared with the standardised mean difference (SMD), which expresses the difference between groups in standard deviation units and, unlike a p-value, does not depend on sample size. An absolute SMD below 0.1 indicates a negligible difference. All SMDs were below 0.1: patients with and without a post-fitting speech test had similar age, sex, hearing aid experience, perceived benefit and need category. This supports the view that the analyses of speech perception are representative of the whole sample for these characteristics. The two groups may still differ on variables that were not available in the dataset, such as the centre or the degree of hearing loss.

**Table S1.** Comparison of patients with and without a post-fitting speech test.

| Variable | With speech test | Without speech test | SMD |
|---|---|---|---|
| n | 11,447 | 13,123 | |
| Age, years, mean (SD) | 75.9 (11.5) | 76.0 (12.2) | −0.009 |
| Women, n (%) | 5,978 (52.2) | 6,997 (53.3) | −0.022 |
| First-time users, n (%) | 7,244 (64.0) | 7,825 (60.6) | 0.070 |
| Degree of Improvement, mean (SD) | 2.24 (0.72) | 2.26 (0.74) | −0.036 |
| Final Ability, mean (SD) | 2.17 (0.72) | 2.20 (0.75) | −0.031 |
| Need classified as “Other”, n (%) | 2,838 (24.8) | 3,173 (24.2) | 0.014 |

A post-fitting speech test was defined as an available post-fitting SIQ or SIN. Percentages are calculated on patients with available data. SMD: standardised mean difference (with minus without); $|SMD| < 0.1$ indicates a negligible difference. SD: standard deviation.

#### *Perceived benefit and speech perception*

*Correlation matrix*

Figure S1 shows the Pearson correlations between age, the two COSI subscores and the four speech tests (SIQ and SIN, before and after fitting). Each cell gives the correlation between the variable of its row and the variable of its column; red indicates a positive correlation, blue a negative correlation, and darker colours stronger correlations. Each correlation was computed on the patients with data for both variables (pairwise n from 614 to 24,267). Higher SIQ and SIN values indicate poorer speech perception.

The speech tests were moderately to strongly correlated with each other (r = 0.45 to 0.69), and age was moderately correlated with all of them (r = 0.31 to 0.35). The two COSI subscores were strongly correlated with each other (r = 0.73) but only weakly correlated with the speech tests (r = −0.15 to 0.06) and with age (r = −0.05 and −0.06). Correlations between perceived benefit and pre-fitting speech tests were close to zero (r = 0.01 to 0.06).

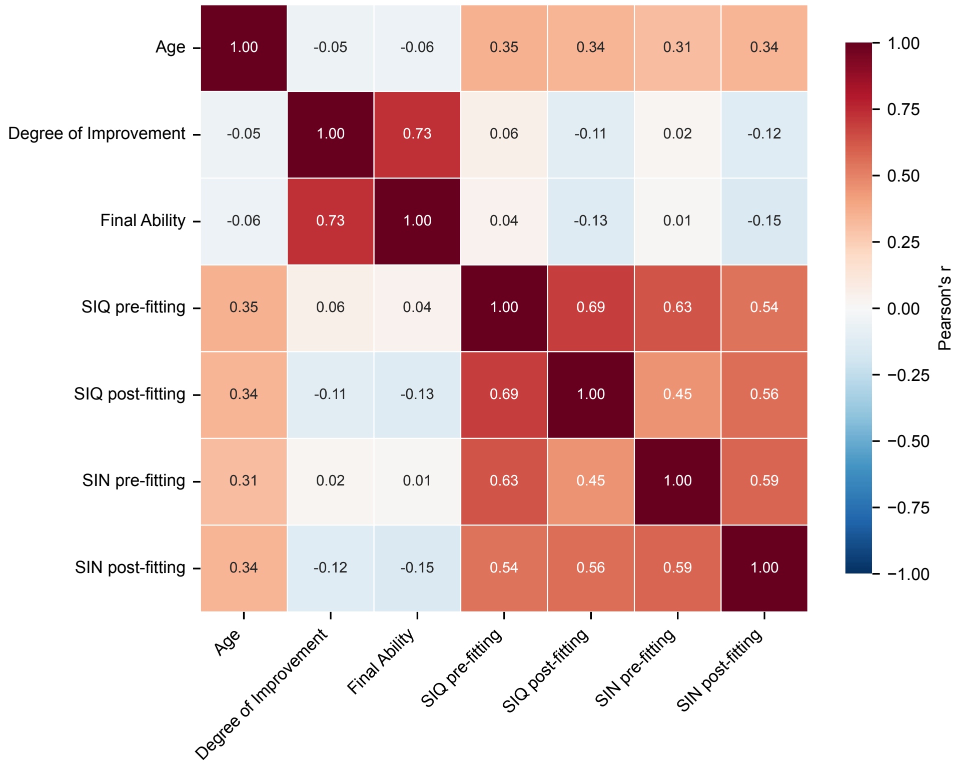


**Figure S1.** Pearson correlations between age, COSI subscores (Degree of Improvement, Final Ability) and speech perception before and after fitting (SIQ: speech in quiet, dB HL; SIN: speech in noise, dB SNR). Each correlation was computed on the patients with data for both variables (pairwise n from 615 to 24,343). Higher SIQ and SIN values indicate poorer speech perception.

*Sensitivity analyses*

Table S2 tests whether the correlations between perceived benefit and speech perception depend on two choices made in the main analysis: which questionnaire was kept for each patient, and how long after the first COSI interview the questionnaire was completed. Each column repeats the correlations of the main analysis in a different sample. Higher SIQ and SIN values indicate poorer speech perception, so a negative correlation means that better speech perception was associated with more perceived benefit. For change scores (pre-fitting minus post-fitting), a positive correlation means that a larger measured improvement was associated with more perceived benefit.

*Main analysis.* The earliest eligible questionnaire of each patient (n = 24,570 patients).

*S1: last questionnaire.* The latest instead of the earliest eligible questionnaire of each patient. This tests whether a later questionnaire, completed after a longer use of the hearing aids, changes the results.

*S2: without questionnaires completed less than 30 days after the first COSI interview.* The date of the first COSI interview was known for 4,629 patients (18.8%); 1,408 of them completed the questionnaire less than 30 days after this interview and were removed. Patients with an unknown date were kept.

*S3 and S4: subgroup with a known date of the first COSI interview.* S3 includes all patients of this subgroup and S4 only those who completed the questionnaire 30 days or more after the first interview. This subgroup differed from the rest of the sample (mean age 73.1 vs 76.6 years), so S4 should be compared with S3 (same patients), not with the main analysis.

*Interpretation.* The correlations with post-fitting SIQ and SIN were almost unchanged with the last questionnaire (S1; for example, SIQ $\times$ DI $-0.126$ vs $-0.113$) and after removing questionnaires completed within 30 days (S2; $-0.113$). In the subgroup with a known date, the correlations were weaker than in the whole sample (S3; SIQ $\times$ DI $-0.039$), and removing questionnaires completed within 30 days did not change them meaningfully (S4; $-0.017$, with overlapping confidence intervals). The weaker correlations are therefore a feature of this subgroup, not an effect of time since the first interview. All patients with both pre- and post-fitting speech tests belonged to this subgroup, so the change-score correlations are identical in the main analysis and in S3. Overall, the choice of questionnaire and the time since the first COSI interview did not change the conclusions of the main analysis.

**Table S2.** Sensitivity analyses: correlations between perceived benefit and speech perception in five samples. Values are Pearson's r [95% CI] (n).

| Pair | Main analysis<br>Earliest<br>questionnaire | S1<br>Last<br>questionnaire | S2<br>Without delay<br>$<$ 30 days | S3<br>Known delay<br>(all) | S4<br>Known delay<br>$\geq$ 30 days |
|---|---|---|---|---|---|
| SIQ post × DI | −0.113 [−0.133; −0.092]<br>(8,516) | −0.126 [−0.148; −0.104]<br>(7,996) | −0.113 [−0.134; −0.091]<br>(7,953) | −0.039 [−0.087; 0.009]<br>(1,663) | −0.017 [−0.076; 0.042]<br>(1,100) |
| SIN post × DI | −0.121 [−0.144; −0.098]<br>(7,252) | −0.126 [−0.151; −0.101]<br>(5,983) | −0.125 [−0.149; −0.102]<br>(6,667) | −0.048 [−0.091; −0.004]<br>(2,008) | −0.051 [−0.103; 0.000]<br>(1,423) |
| SIQ post × FA | −0.132 [−0.154; −0.110]<br>(7,346) | −0.143 [−0.165; −0.119]<br>(7,006) | −0.132 [−0.155; −0.109]<br>(6,855) | −0.063 [−0.113; −0.012]<br>(1,501) | −0.046 [−0.107; 0.016]<br>(1,010) |
| SIN post × FA | −0.146 [−0.169; −0.122]<br>(6,456) | −0.150 [−0.176; −0.123]<br>(5,346) | −0.151 [−0.176; −0.127]<br>(5,940) | −0.049 [−0.094; −0.003]<br>(1,830) | −0.055 [−0.109; −0.001]<br>(1,314) |
| SIQ change × DI | 0.060 [−0.019; 0.139]<br>(612) | 0.105 [−0.002; 0.210]<br>(336) | 0.095 [−0.007; 0.195]<br>(371) | 0.060 [−0.019; 0.139]<br>(612) | 0.095 [−0.007; 0.195]<br>(371) |
| SIN change × DI | 0.082 [0.034; 0.129]<br>(1,648) | 0.078 [0.011; 0.145]<br>(838) | 0.079 [0.022; 0.135]<br>(1,192) | 0.082 [0.034; 0.129]<br>(1,648) | 0.079 [0.022; 0.135]<br>(1,192) |

DI: Degree of Improvement; FA: Final Ability; SIQ: speech in quiet; SIN: speech in noise. Change = pre-fitting minus post-fitting threshold (positive = improvement). Delay = time between the first COSI interview and the questionnaire, known for 18.8% of patients. S3 and S4 include the same subgroup and should be compared with each other.

#### ***Perceived benefit across patient characteristics***

*Pairwise comparisons between age groups*

Table S3 gives the pairwise comparisons (Tukey's HSD test) that followed the one-way ANOVA of perceived benefit by age group. Each row compares two age groups; the difference is the mean of the older group minus the mean of the younger group, so a negative value means that the older group reported less benefit. Adjusted p-values account for the six comparisons made for each subscore. All differences were small (at most 0.15 points on the −1 to 3 scale). The oldest group (81–99 years) scored lower than all other groups for both subscores.

**Table S3.** Pairwise comparisons of perceived benefit between age groups (Tukey's HSD test).

| Comparison (years) | Difference [95% CI] | Adjusted p |
|---|---|---|
| *Degree of Improvement* | | |
| 51–65 vs 18–50 | −0.02 [−0.09; 0.06] | 0.944 |
| 66–80 vs 18–50 | −0.07 [−0.14; 0.00] | 0.066 |
| 81–99 vs 18–50 | −0.12 [−0.19; −0.05] | <0.001 |
| 66–80 vs 51–65 | −0.05 [−0.09; −0.01] | 0.004 |
| 81–99 vs 51–65 | −0.10 [−0.14; −0.06] | <0.001 |
| 81–99 vs 66–80 | −0.05 [−0.08; −0.02] | <0.001 |
| *Final Ability* | | |
| 51–65 vs 18–50 | −0.06 [−0.14; 0.02] | 0.251 |
| 66–80 vs 18–50 | −0.08 [−0.15; 0.00] | 0.035 |
| 81–99 vs 18–50 | −0.15 [−0.22; −0.07] | <0.001 |
| 66–80 vs 51–65 | −0.02 [−0.06; 0.02] | 0.579 |
| 81–99 vs 51–65 | −0.09 [−0.13; −0.05] | <0.001 |
| 81–99 vs 66–80 | −0.07 [−0.10; −0.04] | <0.001 |

Difference = mean of the older group minus mean of the younger group (scale −1 to 3). CI: confidence interval adjusted for multiple comparisons. Group means and sample sizes: Degree of Improvement 2.33 (n = 776), 2.31 (n = 3,065), 2.26 (n = 11,288) and 2.21 (n = 9,138); Final Ability 2.29 (n = 707), 2.23 (n = 2,798), 2.21 (n = 10,167) and 2.14 (n = 8,195), from the youngest to the oldest group.

#### ***Measurement properties***

*How to read the Bland–Altman plot*

The Bland–Altman plot (Figure S2) shows, for each patient, how much the two COSI subscores agree. Two values are computed from the Degree of Improvement (DI) and the Final Ability (FA) of each patient, both scored from −1 to 3.

*Horizontal axis.* The mean of the two subscores shows the overall level of benefit of the patient. For example, DI = 3 and FA = 2 give a mean of 2.5. Values further to the right indicate more benefit.

*Vertical axis.* The difference DI minus FA shows the gap between the two subscores. A value of 0 means that the patient gave the same score to both questions. A value of +1 means that DI was one point higher than FA (for example, "much better" but "most of the time"); a value of −1 means that FA was one point higher than DI.

*Circles.* Because the subscores take only five values, many patients share the same combination of scores. Each circle therefore represents one combination, and its area is proportional to the number of patients. For example, the largest circle (37% of patients) corresponds to DI = 2 and FA = 2. The circle at a mean of 3 and a difference of 0 (32% of patients) corresponds to patients who gave the highest score to both questions, which reflects the ceiling effect.

*Horizontal lines.* The solid line shows the mean bias: on average, DI was 0.12 points higher than FA. The dashed lines show the 95% limits of agreement (−0.89 to 1.13): for 95% of patients, the difference between DI and FA lies between these two values. The dotted line at 0 shows perfect agreement.

*Interpretation.* The two subscores were identical for three patients out of four (75.6%). When they differed, they almost always differed by one point only (23.3%); a difference of two points or more was rare (1.1%) (Table S4). The mean bias comes from an asymmetry: a DI higher than FA (17.8%) was about three times more frequent than the reverse (5.5%).

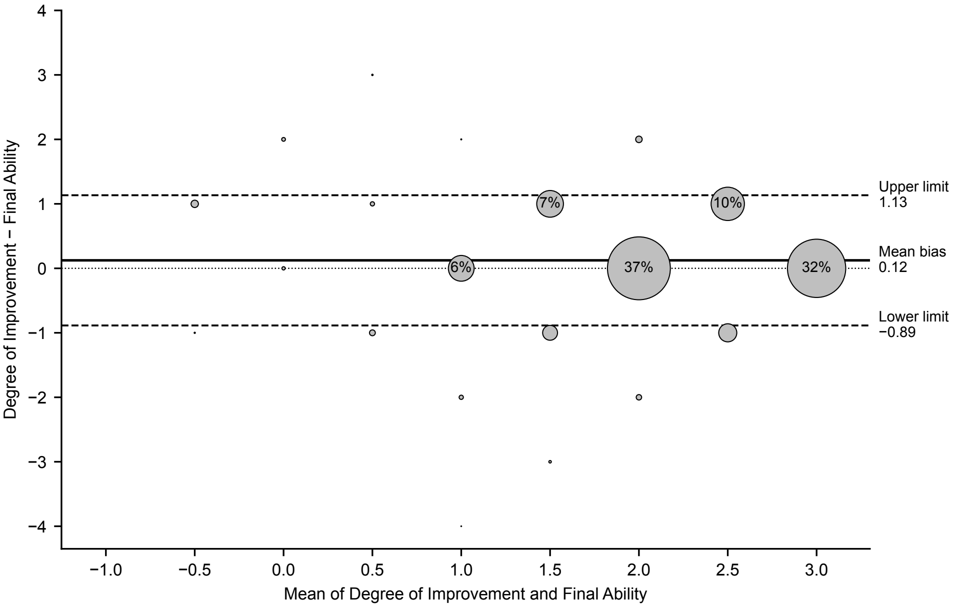


**Figure S2.** Bland–Altman plot of the agreement between the Degree of Improvement and the Final Ability (n = 21,711 patients with both subscores). Each circle represents one combination of scores; its area is proportional to the number of patients, and circles representing at least 5% of patients show this percentage. The solid line shows the mean bias (0.12), the dashed lines the 95% limits of agreement (−0.89 to 1.13), and the dotted line perfect agreement (0).

*Distribution of the difference between Degree of Improvement and Final Ability*

Table S4 gives the exact percentages behind Figure S2. For each patient with both subscores (n = 21,711), the Final Ability was subtracted from the Degree of Improvement. A difference of 0 means that the two subscores were identical; a positive difference means that DI was higher than FA, and a negative difference the reverse.

*Interpretation.* Three patients out of four (75.6%) gave the same score to both questions. Most other patients differed by one point (23.3%), and a difference of two points or more was rare (1.1%). A DI higher than FA (17.8%) was about three times more frequent than the reverse (5.5%); this asymmetry explains the mean bias of 0.12 points shown in Figure S2.

**Table S4.** Distribution of the difference between Degree of Improvement and Final Ability (n = 21,711).

| Difference (DI − FA) | Patients, % |
|---|---|
| −4 | <0.1 |
| −3 | 0.1 |
| −2 | 0.4 |
| −1 | 5.5 |
| 0 | 75.6 |
| +1 | 17.8 |
| +2 | 0.5 |
| +3 | <0.1 |

DI: Degree of Improvement; FA: Final Ability. Both subscores range from −1 to 3. A positive difference means that DI was higher than FA. Percentages may not sum to 100 because of rounding.

### *Coverage of the COSI taxonomy*

#### *Distribution of the first-ranked COSI needs*

Table S5 gives the exact number and percentage of patients in each need category shown in Figure 3A of the main manuscript, with the French label used in the COSI and the classifier. The proportion of "Other" needs is given both as observed and after correction for classifier performance (corrected proportion = observed proportion × precision / recall, with precision 0.83 and recall 0.94 for the "Other" category).

**Table S5.** Distribution of the first-ranked COSI needs (n = 24,570 patients).

| Need category | French label | n | % |
|---|---|---|---|
| Conversation with 1 or 2 in quiet | Conversation à 1 ou 2 dans le silence | 5,250 | 21.4 |
| Conversation with group in noise | Conversation en groupe dans le bruit | 5,231 | 21.3 |
| Television/radio at normal volume | Télévision/radio au volume normal | 3,143 | 12.8 |
| Conversation with group in quiet | Conversation en groupe dans le silence | 2,059 | 8.4 |
| Feel embarrassed or stupid | Se sentir embarrassé ou stupide | 1,708 | 7.0 |
| Church or meeting | Église ou réunion | 733 | 3.0 |
| Unfamiliar speaker on phone | Interlocuteur non familier au téléphone | 273 | 1.1 |
| Increased social contact | Augmenter le contact social | 160 | 0.7 |
| Other (observed) | Autres | 6,013 | 24.5 |
| Other (corrected for classifier performance) | | | 21.6 |

The classifier assigned eight of the predefined COSI categories in this sample; "Other" groups the needs that did not match any of them. Percentages are calculated on all patients and may not sum to 100 because of rounding.

#### *Age and perceived benefit by need category*

Table S6 compares age and perceived benefit across the eight predefined need categories (n = 18,557 patients; "Other" is excluded because it groups heterogeneous needs).

Panel A gives the mean (SD) and sample size of each variable in each category, with the results of the one-way ANOVA and of the Kruskal–Wallis test. Panel B lists the pairs of categories that differed significantly after Tukey's correction for multiple comparisons; for each pair, the difference is the mean of the higher category minus the mean of the lower category.

Age differed most across categories ($\eta^2 = 0.032$): patients whose first need was "Church or meeting" were 7 to 11 years younger than patients in all other categories. Perceived benefit also differed, but the effects were negligible ($\eta^2 = 0.006$ for Degree of Improvement and 0.005 for Final Ability): the largest difference between two categories was 0.23 points on the −1 to 3 scale.

**Table S6.** Age and perceived benefit by need category (predefined categories only, n = 18,557). Panel A: descriptive statistics and one-way ANOVA.

| Need category | Age, years | Degree of Improvement | Final Ability |
|---|---|---|---|
| Conversation with 1 or 2 in quiet | 77.6 (11.4); 5,235 | 2.27 (0.68); 5,204 | 2.20 (0.67); 4,715 |
| Conversation with group in noise | 75.2 (11.4); 5,219 | 2.20 (0.76); 5,175 | 2.15 (0.77); 4,707 |
| Television/radio at normal volume | 78.1 (9.8); 3,134 | 2.18 (0.73); 3,115 | 2.09 (0.77); 2,814 |
| Conversation with group in quiet | 76.5 (12.4); 2,048 | 2.34 (0.68); 2,049 | 2.14 (0.66); 1,832 |
| Feel embarrassed or stupid | 75.5 (11.4); 1,700 | 2.30 (0.69); 1,699 | 2.26 (0.69); 1,506 |
| Church or meeting | 67.9 (13.6); 730 | 2.17 (0.75); 727 | 2.10 (0.75); 636 |
| Unfamiliar speaker on phone | 78.4 (11.2); 273 | 2.10 (0.81); 272 | 2.12 (0.74); 237 |
| Increased social contact | 75.7 (11.5); 160 | 2.26 (0.68); 159 | 2.09 (0.76); 148 |
| ANOVA | F(7, 18,491) = 88.04 | F(7, 18,392) = 17.16 | F(7, 16,587) = 12.07 |
| $\eta^2$ | 0.032 | 0.006 | 0.005 |
| Kruskal–Wallis H | 538.2 | 101.5 | 74.3 |

Values are mean (SD); n. Age is in years; Degree of Improvement and Final Ability range from −1 to 3. All ANOVA and Kruskal–Wallis tests: $p < 0.001$.

**Table S6.** (continued) Panel B: significant pairwise comparisons between need categories (Tukey's HSD test).

| Higher category vs lower category | Difference [95% CI] | Adjusted p |
|---|---|---|
| *Age, years* (16 of 28 pairs) | | |
| Unfamiliar speaker on phone vs Church or meeting | 10.6 [8.1; 13.0] | <0.001 |
| Television/radio at normal volume vs Church or meeting | 10.3 [8.8; 11.7] | <0.001 |
| Conversation with 1 or 2 in quiet vs Church or meeting | 9.7 [8.3; 11.1] | <0.001 |
| Conversation with group in quiet vs Church or meeting | 8.6 [7.1; 10.1] | <0.001 |
| Increased social contact vs Church or meeting | 7.8 [4.8; 10.8] | <0.001 |
| Feel embarrassed or stupid vs Church or meeting | 7.6 [6.1; 9.2] | <0.001 |
| Conversation with group in noise vs Church or meeting | 7.3 [6.0; 8.7] | <0.001 |
| Unfamiliar speaker on phone vs Conversation with group in noise | 3.2 [1.1; 5.4] | <0.001 |
| Television/radio at normal volume vs Conversation with group in noise | 2.9 [2.2; 3.7] | <0.001 |
| Unfamiliar speaker on phone vs Feel embarrassed or stupid | 2.9 [0.7; 5.2] | 0.002 |
| Television/radio at normal volume vs Feel embarrassed or stupid | 2.6 [1.6; 3.7] | <0.001 |
| Conversation with 1 or 2 in quiet vs Conversation with group in noise | 2.4 [1.7; 3.1] | <0.001 |
| Conversation with 1 or 2 in quiet vs Feel embarrassed or stupid | 2.1 [1.1; 3.0] | <0.001 |
| Television/radio at normal volume vs Conversation with group in quiet | 1.7 [0.7; 2.7] | <0.001 |
| Conversation with group in quiet vs Conversation with group in noise | 1.3 [0.4; 2.2] | <0.001 |
| Conversation with 1 or 2 in quiet vs Conversation with group in quiet | 1.1 [0.2; 2.0] | 0.004 |
| *Degree of Improvement* (13 of 28 pairs) | | |
| Conversation with group in quiet vs Unfamiliar speaker on phone | 0.23 [0.09; 0.37] | <0.001 |
| Feel embarrassed or stupid vs Unfamiliar speaker on phone | 0.20 [0.06; 0.34] | <0.001 |
| Conversation with 1 or 2 in quiet vs Unfamiliar speaker on phone | 0.17 [0.03; 0.30] | 0.004 |
| Conversation with group in quiet vs Church or meeting | 0.17 [0.07; 0.26] | <0.001 |
| Conversation with group in quiet vs Television/radio at normal volume | 0.16 [0.10; 0.22] | <0.001 |
| Conversation with group in quiet vs Conversation with group in noise | 0.14 [0.08; 0.20] | <0.001 |
| Feel embarrassed or stupid vs Church or meeting | 0.13 [0.04; 0.23] | <0.001 |
| Feel embarrassed or stupid vs Television/radio at normal volume | 0.12 [0.06; 0.19] | <0.001 |
| Feel embarrassed or stupid vs Conversation with group in noise | 0.11 [0.05; 0.17] | <0.001 |
| Conversation with 1 or 2 in quiet vs Church or meeting | 0.10 [0.02; 0.19] | 0.009 |
| Conversation with 1 or 2 in quiet vs Television/radio at normal volume | 0.09 [0.04; 0.14] | <0.001 |
| Conversation with 1 or 2 in quiet vs Conversation with group in noise | 0.08 [0.03; 0.12] | <0.001 |
| Conversation with group in quiet vs Conversation with 1 or 2 in quiet | 0.06 [0.01; 0.12] | 0.013 |
| *Final Ability* (9 of 28 pairs) | | |
| Feel embarrassed or stupid vs Television/radio at normal volume | 0.18 [0.11; 0.25] | <0.001 |
| Feel embarrassed or stupid vs Church or meeting | 0.16 [0.06; 0.26] | <0.001 |
| Feel embarrassed or stupid vs Conversation with group in noise | 0.12 [0.05; 0.18] | <0.001 |
| Feel embarrassed or stupid vs Conversation with group in quiet | 0.12 [0.05; 0.20] | <0.001 |
| Conversation with 1 or 2 in quiet vs Television/radio at normal volume | 0.11 [0.06; 0.17] | <0.001 |
| Conversation with 1 or 2 in quiet vs Church or meeting | 0.10 [0.00; 0.19] | 0.032 |
| Conversation with 1 or 2 in quiet vs Conversation with group in noise | 0.06 [0.01; 0.10] | 0.005 |
| Conversation with 1 or 2 in quiet vs Conversation with group in quiet | 0.06 [0.00; 0.12] | 0.048 |
| Conversation with group in noise vs Television/radio at normal volume | 0.06 [0.01; 0.11] | 0.013 |

Only significant pairs are shown (adjusted $p < 0.05$). Difference = mean of the higher category minus mean of the lower category; CI adjusted for multiple comparisons.

#### *Sex by need category*

Table S7 shows how often women and men reported each first-ranked need. The $\chi^2$ test was run on the eight predefined categories (main analysis) and repeated with "Other" included. For each category, the adjusted standardised (Haberman) residual compares the observed number of women with the number expected if sex and need category were independent: a positive residual means that women reported this need more often than expected, a negative residual that men reported it more often. Residuals are given for women only, because the residual for men has the same value with the opposite sign. p-values were corrected for multiple testing with the Benjamini–Hochberg procedure.

The association between sex and need category was weak (Cramér's V = 0.09).

Women more often reported the telephone with an unfamiliar speaker (73.3% women), feeling embarrassed or stupid (59.4%), television or radio (55.7%) and conversation with a group in quiet (55.9%), whereas men more often reported conversation with a group in noise and conversation with one or two people in quiet. Two categories showed no difference ("Church or meeting" and "Increased social contact"). Including "Other" gave the same pattern (V = 0.08).

**Table S7.** Sex by need category: counts and adjusted standardised residuals.

| | | | Predefined categories | | Including "Other" | |
|---|---|---|---|---|---|---|
| Need category | Women, n (%) | Men, n | Residual | p (FDR) | Residual | p (FDR) |
| Conversation with 1 or 2 in quiet | 2,607 (49.7) | 2,643 | −4.6 | <0.001 | −5.2 | <0.001 |
| Conversation with group in noise | 2,544 (48.6) | 2,687 | −6.4 | <0.001 | −6.8 | <0.001 |
| Television/radio at normal volume | 1,751 (55.7) | 1,392 | +4.1 | <0.001 | +3.5 | <0.001 |
| Conversation with group in quiet | 1,150 (55.9) | 908 | +3.4 | <0.001 | +2.9 | 0.005 |
| Feel embarrassed or stupid | 1,014 (59.4) | 694 | +6.1 | <0.001 | +5.6 | <0.001 |
| Church or meeting | 367 (50.1) | 366 | −1.3 | 0.232 | −1.5 | 0.147 |
| Unfamiliar speaker on phone | 200 (73.3) | 73 | +7.0 | <0.001 | +6.8 | <0.001 |
| Increased social contact | 85 (53.1) | 75 | +0.2 | 0.848 | +0.1 | 0.936 |
| Other | 3,257 (54.2) | 2,756 | – | – | +2.4 | 0.020 |
| $\chi^2$ (df); Cramér's V | | | 151.9 (7); 0.09 | | 157.9 (8); 0.08 | |
| n | | | 18,556 | | 24,569 | |

Residual: adjusted standardised (Haberman) residual for women; the residual for men has the same value with the opposite sign. A positive residual means that women reported this need more often than expected. p (FDR): p-value corrected with the Benjamini–Hochberg false discovery rate procedure. Both $\chi^2$ tests: $p < 0.001$. One patient with missing sex was excluded.

*Lexicon of the themes within "Other"*

Table S8 lists the lexical patterns used to tag the first-ranked "Other" responses (n = 6,013). Patterns were matched, case- and accent-insensitive, on the normalised French text; a response received a theme when it matched at least one pattern of that theme, and could receive several themes. The patterns are shown in simplified form (word stems and expressions); the exact regular expressions are in the script of the public repository (`https://github.com/mcampi111/COSI`).

**Table S8.** Themes within the “Other” responses: lexical patterns and prevalence (n = 6,013).

| Theme | French patterns (simplified) | n (%) |
|---|---|---|
| Device discretion, aesthetics and format | discre-, invisible, intra-, esthét-, petit, rechargeab-, pile, qui (ne) se voi-, contour | 882 (14.7) |
| Tinnitus management | acouph-, acoufe-, accouph-, bourdonnement, sifflement, tinnitus | 771 (12.8) |
| Price and administrative | 100% santé, RAC 0, reste à charge, MDPH, MDA, CMU, CSS, devis, renouvel-, rempla-, mutuelle, prix, cher, abordable | 632 (10.5) |
| Weak, distant or masked voices | voix faible, voix basse, chuchot-, parle (bas / doucement / faible), de loin, éloign-, à distance, lointain, masque, lecture labiale, sans voir, n'articul- | 555 (9.2) |
| Laterality, stereo and localisation | stéréo, binaural, rééquilibr-, localis-, cône d'ombre, CROS, BAHA, hémichamp, côté (droit / gauche), deux oreilles, d'où vient | 285 (4.7) |
| Paediatric or school | enfant, fils, fille, classe, école, collège, lycée, amphi, cours, maîtresse, professeur, orthophon-, implant cochléaire, apprentissage, langage, diction, oralité, micro HF, Roger | 212 (3.5) |
| Environmental safety (cars, alarms) | sécurité, danger, alerte, alarme, détecteur, sonnette, sonnerie, voiture, train, métro, vélo, conduit-, permis, poids lourd, GPS, signal | 100 (1.7) |
| Music, fine perception and nature | musiqu-, piano, saxophone, cornemuse, cor de chasse, flûte, chorale, chant, instrument, note, violon, contrebasse, chef de chœur, oiseau, cigale, nature | 93 (1.6) |
| Theatre, cinema and lectures | théâtre, cinéma, spectacle, conférence, amphi | 79 (1.3) |
| Hyperacusis, listening fatigue and cognition | hyperacous-, bruit(s) fort(s), son(s) fort(s), intoléran-, fatigu-, concentr-, mémoire, cognit-, attention, reposer le cerveau, agress- | 50 (0.8) |
| Vertigo, balance and pathology | vertige, Ménière, équilibre postural, cholestéatome, otospongiose, neurinome, surdité brusque, surdité brutale, congénital, cophose | 33 (0.6) |

A hyphen after a stem means that any ending was accepted (for example, acouph- matches acouphène and acouphènes). Percentages are calculated on the 6,013 first-ranked “Other” responses of the final sample and do not sum to 100 because a response could receive several themes.

### Conclusions
